# Women’s empowerment and publicly-funded health insurance in India: associations with PM-JAY verification, outpatient service use, and out-of-pocket spending across six states

**DOI:** 10.64898/2026.10.19.26365973

**Authors:** Swati Srivastava, Susanne Ziegler, Divya Parmar, Christoph Strupat, Manuela De Allegri

**Affiliations:** Heidelberg Institute of Global Health, Medical Faculty and University Hospital, Heidelberg University, Im Neuenheimer Feld 130.3, 69120 Heidelberg, Germany; Public Health Foundation of India, Saidulajab Village, Block H, Saket, New Delhi, Delhi 110017, India; Deutsche Gesellschaft für Internationale Zusammenarbeit (GIZ) GmbH, Friedrich-Ebert-Allee 32+36, 53113 Bonn, Germany; Department of Population Health Sciences, School of Life Course and Population Sciences, King’s College London, Weston Education Centre, Cutcombe Road, London SE5 9RJ, United Kingdom; German Institute of Development and Sustainability (IDOS) gGmbH, Tulpenfeld 6, 53113 Bonn, Germany

**Keywords:** Empowerment, women, PM-JAY, verification, outpatient care, out-of-pocket expenditure

## Abstract

**Objectives:** Women’s empowerment is a key determinant of health and well-being and may shape how women access and benefit from publicly-funded health insurance (PFHI). We aimed to examine whether different levels of women’s empowerment were associated with 1) verification in India’s PFHI Pradhan Mantri Jan Arogya Yojana (PM-JAY), 2) outpatient care utilization, and 3) resultant out-of-pocket expenditure.

**Design:** We conducted a cross-sectional observational study. We analyzed data from a household survey, created a composite empowerment index using structural equation models, and used multivariate regression models to test associations between women’s empowerment and the outcome variables.

**Setting:** The study was conducted between December 2019 and February 2020 among PM-JAY eligible households in six Indian states (Bihar, Chhattisgarh, Gujarat, Meghalaya, Tamil Nadu, and Uttar Pradesh).

**Participants:** 14,378 women across the six states.

**Results:** Three main findings emerged: First, high levels of women’s empowerment were associated with higher odds of completing PM-JAY verification (AOR: 1.22, 95% CI: 1.09-1.36). Second, women’s empowerment was not associated with outpatient service utilization. And third, either medium levels of empowerment (coef = -0.49, p = 0.020) or PM-JAY verification (coef = -0.37, p = 0.034) was associated with lower OOPE for outpatient services. Women who had both, PM-JAY verification and medium levels of empowerment, experienced higher OOPE (coef = 0.45, p = 0.058).

**Conclusions:** Women’s empowerment is an important factor related with PM-JAY verification, but not with outpatient service use. OOPE varied depending on whether women were both empowered and PM-JAY verified or not. Our results suggest that institutional and structural barriers may play a greater role in healthcare access than individual factors pertaining to empowerment alone. Achieving equitable access therefore requires gender-responsive systems that reduce administrative barriers and support women in navigating the healthcare pathway. Our findings highlight the importance of integrating outpatient care into PM-JAY to increase financial protection, streamlining verification processes to improve women’s engagement with PM-JAY and the broader health system, and advancing health financing reforms that better support women.

**STRENGTHS AND LIMITATIONS OF THIS STUDY:**

- We used a large cross-sectional household survey representative of PM-JAY eligible households from districts across six Indian states.
- We constructed a composite empowerment index based on established theoretical domains of empowerment and estimated it using structural equation modelling.
- Although outpatient care is not covered under PM-JAY, we focused specifically on outpatient services as little is known about the income and substitution effects of PM-JAY coverage on outpatient service utilization.
- Sampling lists used for the study derived from the official PM-JAY targeting database from 2011; many households have since changed social status and were untraceable in urban areas due to incorrect identifiers. This restricted data collection to primarily rural and semi-urban (as opposed to urban) areas.
- A reliance on self-reported data may have introduced selection and recall biases.

## INTRODUCTION

Empowered women and girls are central to achieving just, inclusive societies in which people can realize their potential and live healthy, socially productive lives. This idea is reflected in Sustainable Development Goal 5 on gender equality and Goal 3 on health and well-being, which includes universal health coverage (UHC)[1,2]. Empowerment is widely conceptualized through three linked dimensions: resources, agency, and achievements[3].

Resources refer to the material, social, and human conditions that enable action; agency refers to women’s ability to define goals and act on them; and achievements refer to the outcomes that follow, such as improved health or greater control over life circumstances[3]. Further, empowerment is both a process and an outcome: it increases women’s capacity to make meaningful life choices in constrained settings, and in doing so helps challenge social, economic, political, and institutional inequalities[3]. Women’s empowerment generates a range of positive outcomes. At the individual level, it strengthens women’s economic assets, enhances their decision-making power, and improves self-perceptions[4]. It is associated with better health outcomes for women themselves, as well as for their children and families[5]. At the societal level, women’s empowerment contributes to broader social and political mobilization, enabling women to influence and reshape gendered institutions and systems[6].

Publicly-funded health insurance (PFHI) schemes, in which monetary contributions for poorer or disadvantaged populations are borne by the state, are increasingly being used to improve healthcare utilization, financial protection, and overall health outcomes, aligned with the global impetus on attaining UHC[7]. In the context of empowerment, PFHI schemes engender all three domains of resources, agency, and achievements. PHFI increase access to resources through both income effects (by reducing out-of-pocket expenditure [OOPE], effective income is increased, which can increase demand for healthcare, even other healthcare services not covered by the service package, such as outpatient services) and through substitution effects (lower OOPE implies that healthcare utilization becomes cheaper compared to other goods, and users substitute toward more healthcare)[8–11]. PFHI has been observed to increase agency by enabling women to make healthcare decisions without having to negotiate household resources[12]. Yet, we can postulate theoretically that the relationship between empowerment and PFHI may be bi-directional: already empowered women may theoretically engage more with PFHI services[5,13,14], and as a result become more empowered through better health and greater economic capital through cost- and time-savings and increased labor productivity, and therefore have greater control over life choices and agency, such as for healthcare decisions. While there is limited evidence from PHFI, there is ample evidence to show that social protection programs positively affect these pathways[15–18]. In order to achieve these gains, women’s engagement with and navigation through the PFHI system is critical: first, through formal registration or enrolment into the scheme, second, through utilization of services, and third, from the resulting financial protection. Empowered women may engage more across this cascade due to greater health literacy, higher bargaining power within the family, and thus exercise greater control over their own health choices and those of their children.

Still, the relationship between women’s empowerment and health insurance has been shown to be highly context-dependent. In Rwanda, J. Kawuki et al. (2022) found paradoxically negative associations between women’s empowerment indicators and health insurance coverage among women aged 15–49 years: women with high economic empowerment and decision-making power were less likely to have health insurance coverage than women with no economic empowerment or decision-making power[19]. This may have been because empowered women were wealthier and had a lower perceived need for health insurance.

Conversely, in Ethiopia, L. Vértesy & G. Namomsa (2023) reported that community-based health insurance positively fosters women’s health and economic empowerment by reducing OOPE for health and increasing decision-making power[20]. Similarly, A. Seid & M. Ahmed (2021) found that among Ethiopian mothers, health insurance enrolment was associated with a higher likelihood of antenatal care utilization and skilled birth attendance.

In India, the Pradhan Mantri Jan Arogya Yojana (PM-JAY) PFHI scheme was launched in 2018 to provide inpatient coverage to 500 million socio-economically disadvantaged Indians. Existing research on PM-JAY has mainly focused on verification, utilization of inpatient services, and related OOPE. One study found that PM-JAY verification was associated with lower OOPE and catastrophic health expenditure for inpatient services used in private facilities[21], whereas others found that verification had no effect on outpatient OOPE and reported higher OOPE for service obtained in private facilities[22,23]. Although some research has examined women’s engagement with the scheme[24], little is known about how empowerment of women is associated with such engagement. More broadly, existing evidence from India largely focuses on PM-JAY’s predecessor, RSBY (Rashtrya Swasthya Bima Yojana), and some state schemes, showing that women’s use of RSBY was constrained by household and programmatic barriers: enrolment in and utilization of PFHI have been found to be consistently lower among women compared to men[25–27], possibly as a result of being less aware of the scheme and their entitlements[28,29]. However, other research has reported higher enrolment among households headed by women[30–32].

Our analysis focuses specifically on outpatient services, an area that remains largely unexplored in the context of PM-JAY, as most existing research has concentrated on inpatient care[21–23]. Although outpatient care is not covered under PM-JAY, outpatient and inpatient services are intrinsically linked along the continuum of healthcare utilization. Theoretically, we hypothesize that by increasing access to resources and the income and substitution effects described earlier, PM-JAY can affect outpatient utilization. We therefore seek to examine whether women of varying empowerment levels with PM-JAY coverage use outpatient services differently from those without coverage. Specific objectives include assessing whether women’s empowerment is associated with 1) verification in PM-JAY; 2) utilization of outpatient services; and 3) the resultant OOPE.

## MATERIALS AND METHODS

### Study setting

PM-JAY is India’s flagship PFHI scheme aimed at improving financial protection and access to healthcare among socio-economically vulnerable populations. The scheme provides annual health insurance coverage of up to 500,000 INR (Indian Rupees) per family for secondary and tertiary inpatient care to approximately 500 million individuals identified primarily through deprivation and occupational criteria based on the Socio-Economic Caste Census (SECC) 2011. PM-JAY is an entitlement-based scheme. Although there are no requirements for renewing enrolment status and paying premiums, all eligible individuals need to undergo a verification procedure at PM-JAY-empaneled hospitals or through community-level verification centers in lieu of complex enrolment procedures which may require payments or periodic renewals. Such enrolment procedures have been found to deter uptake and use of PFHI schemes[33]. While PM-JAY covers a broad range of inpatient procedures, including hospitalization, diagnostics, and pre- and post-hospitalization expenses, outpatient services and most outpatient medicines are not included within the benefit package.

### Data sources and sampling

The study used data from a cross-sectional household survey conducted between December 2019 and February 2020 in six Indian states: Bihar, Chhattisgarh, Gujarat, Meghalaya, Tamil Nadu, and Uttar Pradesh. The states, and two to three districts within each state, were purposively selected in consultation with Indian government officials and the study funders. This approach was intended to capture India’s geographical diversity and the different implementation models used for PM-JAY, including administration through autonomous agencies, insurance companies, or a combination of both. Villages within the selected districts and participating households were then randomly selected. The sampling strategy is described in detail in the study protocol[34].

The survey used a pre-tested questionnaire that was translated into local languages and administered by trained interviewers fluent in these languages. The questionnaire covered several domains, including health insurance literacy, access to healthcare, outpatient and inpatient service utilization and associated OOPE, as well as awareness of PM-JAY. For this study, we used the survey modules on participants’ socio-economic and demographic characteristics, as well as outpatient service utilization and associated OOPE. We also used a module designed to assess women’s experience with PM-JAY, their perceived roles in household decision-making and their empowerment. Questions within this module were based on validated empowerment and decision-making questions asked in the Demographic and Health Surveys[35]. This module was administered to any adult woman aged 15 years or older who was present at the time of the survey, with one woman interviewed per household. This resulted in a study sample of 14,378 women across the six states.

### Variables and their measurement

#### Measurement of empowerment

We constructed a composite variable reflecting women’s empowerment, based on existing evidence on the different theoretical conceptualizations and domains of women’s empowerment: economic, social, political, psychological, and physical[36–38]. Economic empowerment refers to women’s power and agency within all economic domains; social empowerment refers to women’s abilities to exercise control over non-economic decision making within households; political empowerment encompasses participation in legal rights and political participation; and psychological empowerment describes the ability to make choices and act upon them[39]. Physical empowerment refers to women’s bodily integrity and freedom from physical harm and violence[40]. We mapped available indicators in our dataset to these domains and found 18 items relating to women’s social decision-making (4 items), necessity for permission to go to places (4 items), perception about problems for seeking healthcare (6 items) and economic empowerment (4 items); these are presented in Table 1 below.

**Table 1:**
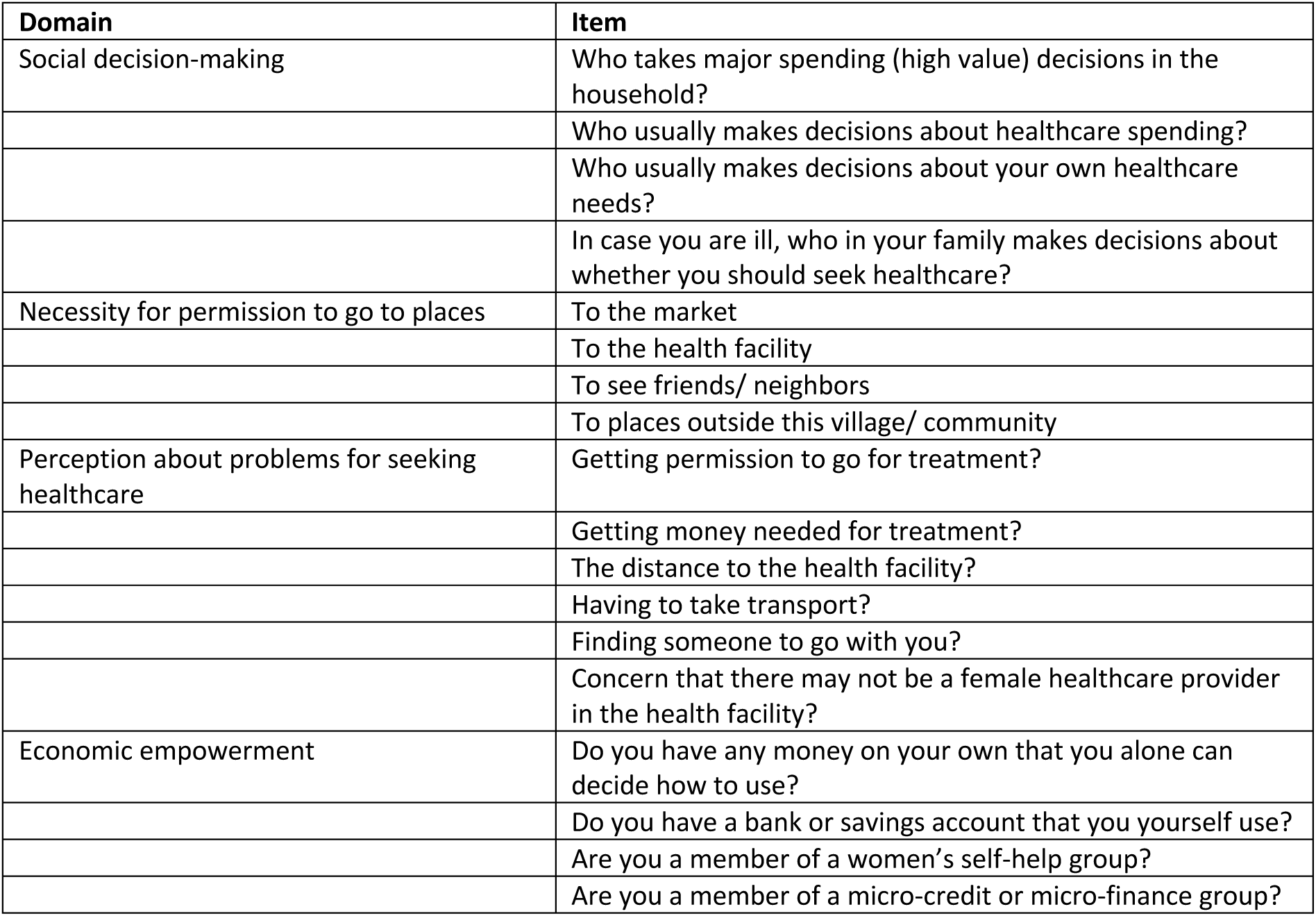
Domains of women’s empowerment and relevant items in our dataset.

First, items were cleaned by checking for missing values and coding direction. There were no missing values, and items were recoded to a binary scale: 0 (no or low empowerment) or 1 (high empowerment), depending upon whether the women alone or woman and husband/partner made decisions (=1) or whether the husband or other family members made decisions on behalf of the woman (=0). Second, the measurement model for the composite empowerment variable was estimated. Within this, we first estimated summary statistics to understand the skewness and kurtosis of the distribution of the items, and examined the correlation matrix to understand which items were correlated. We then performed exploratory factor analysis, using a likelihood ratio test (LRT) to test four and five factor solutions. We found that a four-factor solution had a higher chi-squared value and better fit. Three items were dropped based on low loadings (<0.5) or cross-loadings suggested by modification indices, retaining 15 of the 18 items. These four factors corresponded to the domains of decision-making about self and family (L1), independence in mobility decisions (L2), independence in treatment decisions (L3), and financial empowerment (L4). Third, we performed confirmatory factor analysis using structural equation modelling to estimate a composite score for each respondent. The model was specified with 15 items loading on four latent factors: L1 onto women_spend_dec (λ=0.786, p<0.001), women_healt_dec (λ=0.876, p<0.001), women_ownhealt_dec (λ=0.903, p<0.001), and women_seekhealt_dec (λ=0.848, p<0.001); L2 onto women_places_mrkt (λ=0.767, p<0.001), women_places_hosp (λ=0.808, p<0.001), women_places_friend (λ=0.781, p<0.001), and women_places_out (λ=0.779, p<0.001); L3 onto women_treat_money (λ=0.685, p<0.001), women_treat_dist (λ=0.822, p<0.001), women_treat_transp (λ=0.791, p<0.001), women_treat_comp (λ=0.680, p<0.001), and women_treat_nofem (λ=0.554, p<0.001); and L4 onto women_selfhelp (λ=0.549, p<0.001) and women_micro (λ=0.805, p<0.001). All latents had unit variance, with factor covariances specified for L1-L2 (ρ=0.304, p<0.001), L1-L3 (ρ=0.092, p<0.001), L1-L4 (ρ=-0.005, p=0.631), L2-L3 (ρ=0.123, p<0.001), L2-L4 (ρ=0.158, p<0.001), and L3-L4 (ρ=-0.051, p<0.001). The model was estimated via maximum likelihood (N=14,378) with a diagonal exogenous covariance structure and standardized latent variables. Model fit was acceptable: χ²(84)=4979.09 (p<0.001), RMSEA=0.064 (90% CI: 0.062–0.065), CFI=0.952, TLI=0.940, SRMR=0.028. Detailed tables of the estimation are available in Supplementary File 1.

The empowerment score was computed as the standardized sum of the four latent factor scores (mean of unstandardized items), with higher values indicating greater empowerment. We tested the internal consistency of the scale through Cronbach’s alpha (scale reliability coefficient: 0.7872; average interitem covariance: 0.042). To estimate the categorical empowerment variable used in the analysis, we divided the empowerment score into tertiles to create the following three categories: no or low empowerment, medium empowerment, and high empowerment.

#### Construction of the outcome variables

To address our research questions, we specified three outcome variables. The binary variable *verification in PM-JAY* indicated whether a woman had undergone the verification process, which was a prerequisite for accessing services under PM-JAY (yes/no). The binary variable *outpatient service utilization* denoted whether a woman had used outpatient services in the preceding 30 days (yes/no). *OOPE for outpatient services* was measured as a continuous variable and captured the expenses associated with such service use over the same period. OOPE included doctor or consultation fees, medicines, diagnostic tests, and other related expenditures, such as transportation and food costs.

We controlled for a range of socio-demographic characteristics in the multivariable models, as shown in Table 2. We acknowledge that several of these characteristics may also be associated with women’s empowerment, our primary exposure of interest. Because these factors were included as covariates for confounding control, we did not separately model their associations with women’s empowerment. An overview of all variables, their measurement, and their distribution in the sample is presented in Table 2.

**Table 2:**
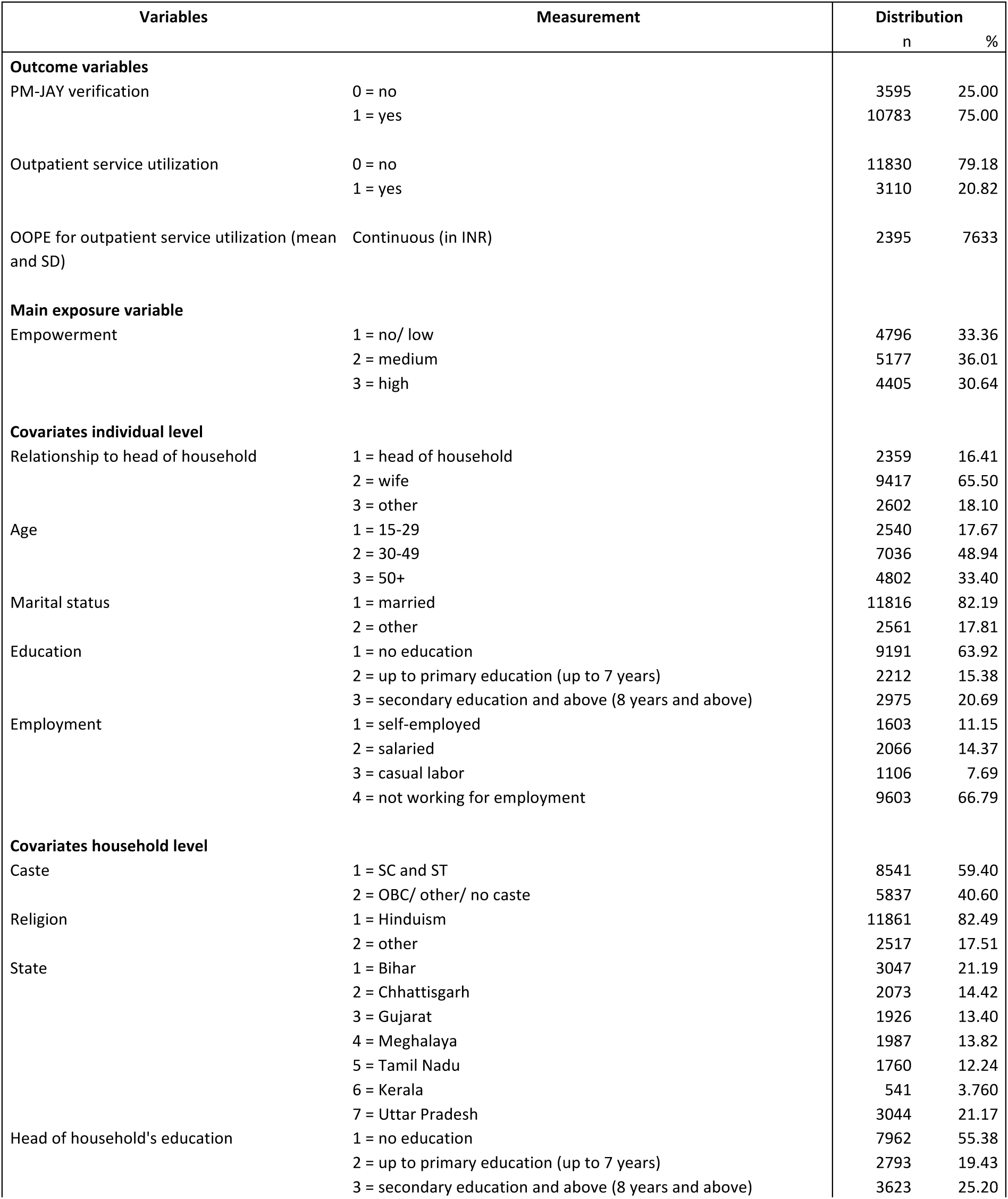
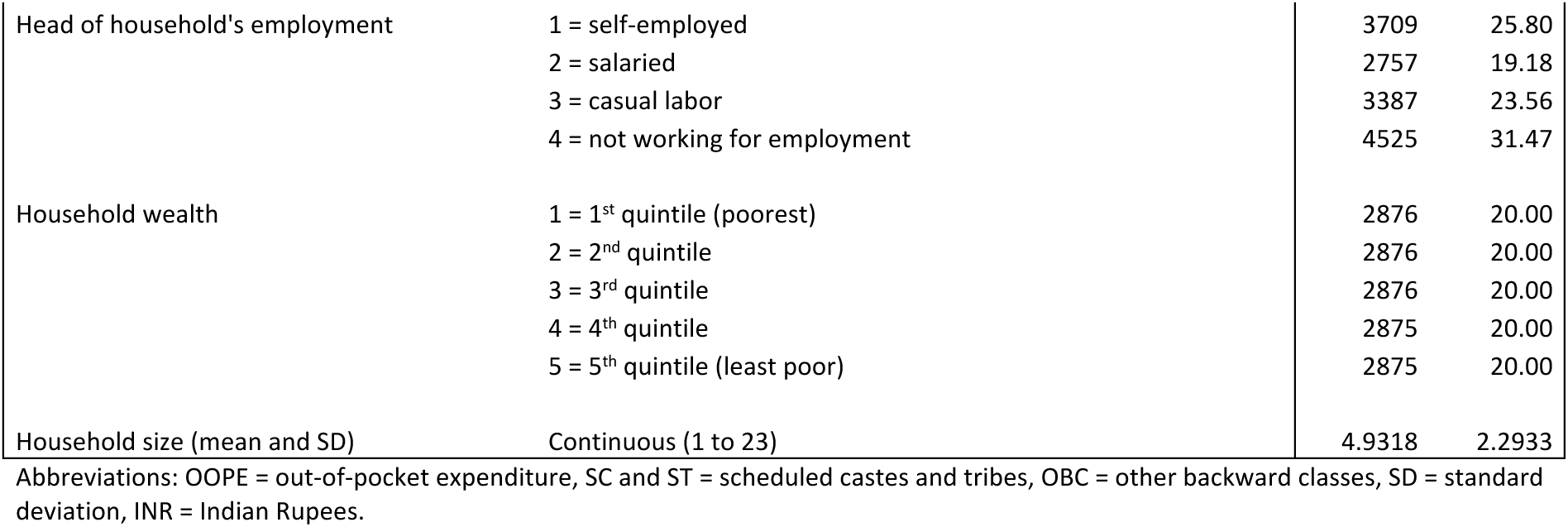
Variables, their measurement, and distribution among women (N=14,378)

### Patient and public involvement

Officials of the Indian central and state governments were involved in the design and implementation of the survey. Results were shared and discussed with them. Informed consent was obtained from the study participants. Patients were not part of the survey or this study.

## ANALYTICAL APPROACH

Given that the primary objective of the study was to examine whether different levels of women’s empowerment were associated with the outcome variables, the categorical empowerment variable (*empow*) was the key explanatory variable of interest. We performed bivariate analyses to examine the distribution of covariates across women who had undergone the verification process and those who had not, as well as between women who used outpatient services and those who did not. We summarized the covariates using means and standard deviations (SDs) and used t-tests and analyses of variance (ANOVA) to assess differences across variable categories. The results of these analyses were then used to inform the following three regression models:

First, for the outcome *verification in PM-JAY*, we fitted a multivariate logistic regression model to estimate the likelihood of a woman having undergone the PM-JAY verification process. The model was specified as follows:

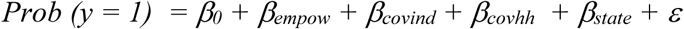

The outcome variable *y* was binary, taking the value 1 if a woman had undergone the verification process and 0 otherwise. *empow* served as the key variable of interest, based on the assumption that verification in PM-JAY was associated with a woman’s level of empowerment. To account for potential confounding, we adjusted for socio-economic and demographic characteristics at both the individual (*covind*) and household (*covhh*) levels as well as state.

Second, for the outcome *utilization*, we estimated a multivariate logistic regression model in which the probability *Prob (y = 1)* — indicating that a woman used outpatient services — was regressed on the same exposure variable and covariates as in the first model. We additionally included *verification* and the interaction term (*verif # empow*) to examine whether, and to what extent, the association between different empowerment levels and outpatient utilization differed depending upon PM-JAY verification status. The outcome was binary, and the model was as follows:

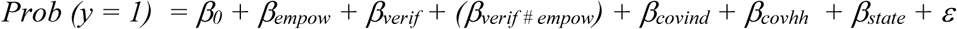

Third, for the outcome *OOPE*, we applied a generalized linear model (GLM) with a gamma distribution and log link function as the outcome was highly skewed. This approach has proven to be well suited to positive right-skewed expenditure data, and it estimates healthcare expenditure more accurately than alternative models[41–45]. The model was expressed as:

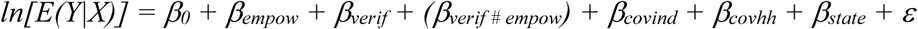

Here, ln[E(Y|X)] denotes the log link function for the outcome *OOPE*. We included the same exposure variable and covariates as in the previous models and also added the interaction term. Additionally, we used the Wald test to assess variable inclusion in each model and the LRT to evaluate interaction terms. The Wald test showed that the selected variables significantly improved the statistical fit across all three models (p < 0.001). The LRT confirmed significance for the GLM regression (p = 0.0958), suggesting that including the interaction term *verif # empow* improved the final model fit. For consistency in data analysis, we retained the interaction term in the logistic regression for the outcome utilization, despite its lack of significance. It is important to note that the OOPE analysis was conditional on reported outpatient service use (and therefore excludes women who were ill but did not seek outpatient services) and those with positive OOPE (92.5% of those who used). All analyses were performed using Stata 16.1[46].

## RESULTS

A total of 14,378 women were included in the analysis. Most women (75%) had completed the PM-JAY verification process, and 20.82% had used outpatient services in the previous 30 days. Among the 3,110 women who used outpatient services in that period, 2,878 (92.5%) incurred some OOPE, with a mean expenditure of 2,395 INR and a standard deviation of 7,633 INR. Based on the empowerment score, 33.36% of women were classified as having no or low empowerment, 36.01% as having medium empowerment, and 30.64% as highly empowered. As PM-JAY targets disadvantaged groups, the sample included a relatively high proportion of women from scheduled castes and tribes (59.4%) and women with no education (63.92%), as well as households headed by persons with no education (55.38%). Most women were married (82.19%) and not engaged in paid work (66.79%). In terms of age, the largest group was women aged 30 to 49 years (48.94%), followed by those aged 50 years and above (33.4%) and those aged 15 to 29 years (17.67%).

In the following and in line with the objective of this study, we describe the results for each outcome, but focus on the main exposure variable *empowerment* and additionally, for outcomes 2 and 3, on *verification in PM-JAY*.

### Empowerment and PM-JAY verification

Table 3, column 1, shows the results of the bivariate analysis for the outcome *verification in PM-JAY*: highly empowered women were more likely to be verified in PM-JAY than women with medium or no/ low empowerment (high: 78.37%, medium: 75.89%, no/ low: 71.64%, p < 0.001).

**Table 3:**
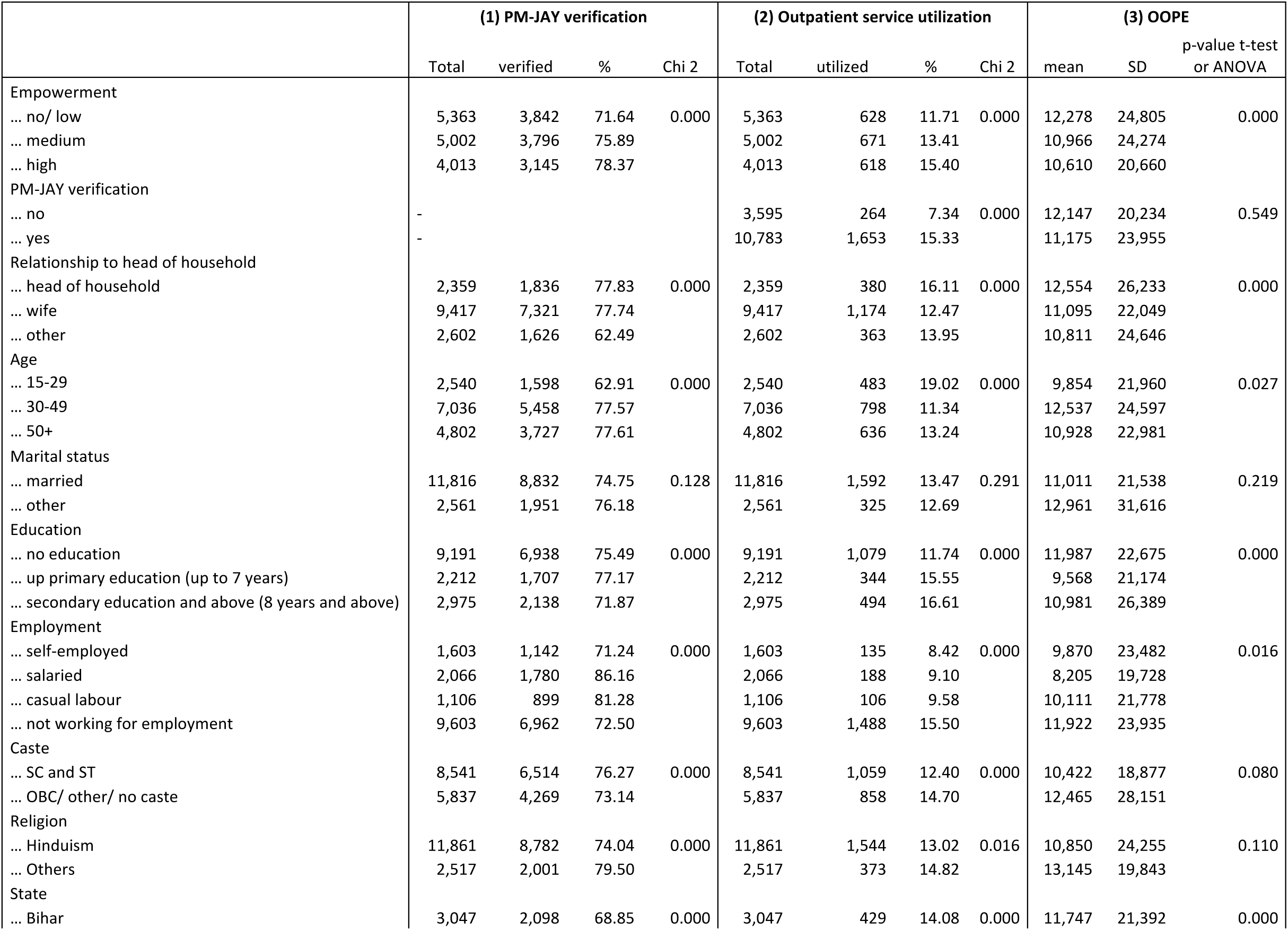
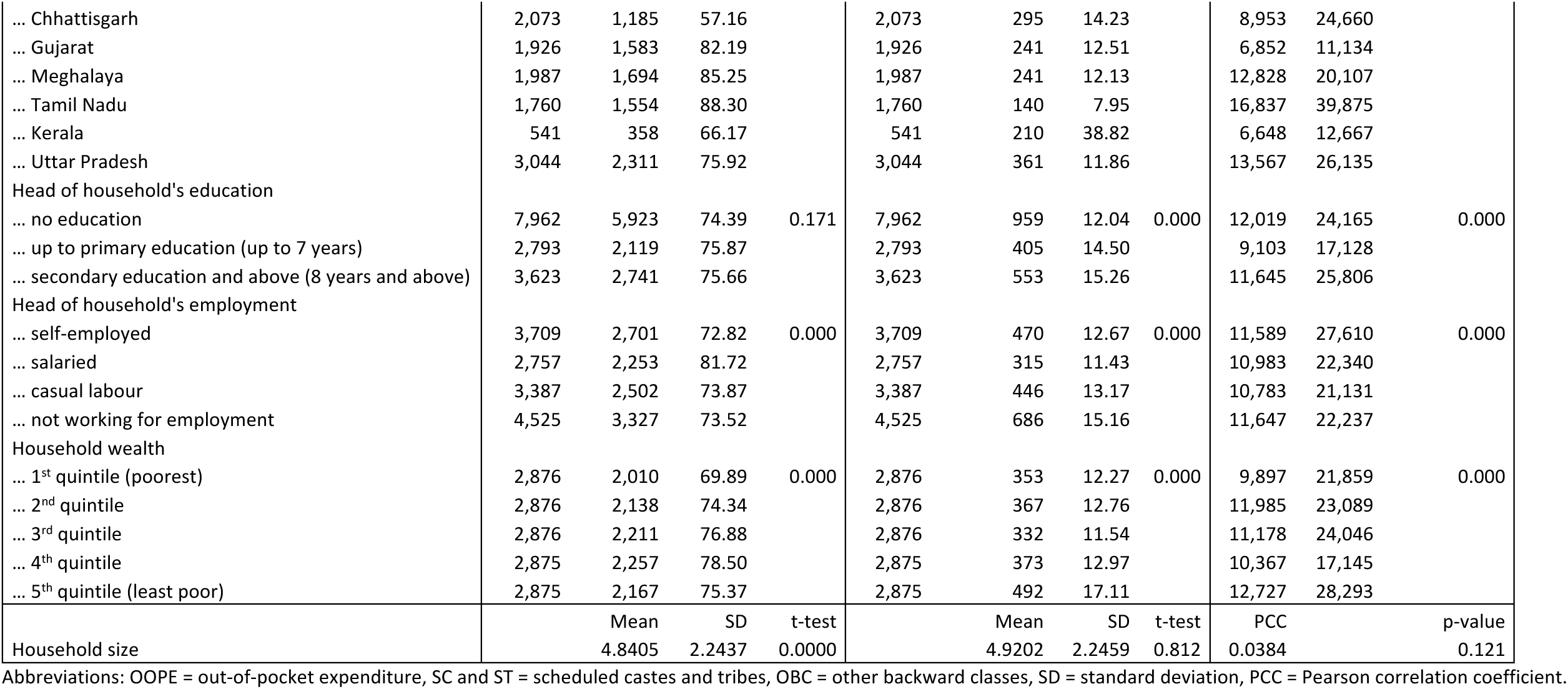
Results of the bivariate analyses.

Multivariate logistic regression in Table 4, column 1, confirmed that women with a high empowerment level had a higher probability of having undergone the PM-JAY verification process (AOR: 1.22, 95% CI: 1.09-1.36) than women with no/ low empowerment.

**Table 4:** Results of the multivariate analyses.

|  | (1) PM-JAY verification |  |  |  |  | (2) Outpatient service utilisation |  |  |  |  | (3) OOPE |  |  |  |  |
| --- | --- | --- | --- | --- | --- | --- | --- | --- | --- | --- | --- | --- | --- | --- | --- |
|  | AOR | 95% CI |  | p-value |  | AOR | 95% CI |  | p-value |  | coef | 95% CI |  | p-value |  |
| Empowerment |  |  |  |  |  |  |  |  |  |  |  |  |  |  |  |
| ... no/ low | 1.00 |  |  |  |  | 1.00 |  |  |  |  | 0.00 |  |  |  |  |
| ... medium | 1.05 | 0.95 | 1.16 | 0.319 |  | 1.00 | 0.81 | 1.24 | 0.971 |  | -0.49 | -0.91 | -0.08 | 0.020 | ** |
| ... high | 1.22 | 1.09 | 1.36 | 0.000 | *** | 1.03 | 0.82 | 1.29 | 0.805 |  | -0.07 | -0.53 | 0.38 | 0.750 |  |
| PM-JAY verification |  |  |  |  |  |  |  |  |  |  |  |  |  |  |  |
| ... no | - |  |  |  |  | 1.00 |  |  |  |  | 0.00 |  |  |  |  |
| ... yes |  |  |  |  |  | 1.51 | 1.27 | 1.80 | 0.000 | *** | -0.37 | -0.70 | -0.03 | 0.034 | ** |
| Verification # empowerment |  |  |  |  |  |  |  |  |  |  |  |  |  |  |  |
| ... verification # medium empowerment | - |  |  |  |  | 0.90 | 0.71 | 1.14 | 0.370 |  | 0.45 | 0.02 | 0.92 | 0.058 | * |
| ... verification # high empowerment |  |  |  |  |  | 0.90 | 0.69 | 1.16 | 0.398 |  | -0.12 | -0.62 | 0.38 | 0.636 |  |
| Relationship to head of household |  |  |  |  |  |  |  |  |  |  |  |  |  |  |  |
| ... head of household | 1.00 |  |  |  |  | 1.00 |  |  |  |  | 0.00 |  |  |  |  |
| ... wife | 1.65 | 1.41 | 1.94 | 0.000 | *** | 0.99 | 0.83 | 1.19 | 0.950 |  | -0.11 | -0.45 | 0.23 | 0.529 |  |
| ... other | 0.87 | 0.73 | 1.03 | 0.100 |  | 0.61 | 0.50 | 0.75 | 0.000 | *** | 0.10 | -0.28 | 0.48 | 0.593 |  |
| Age |  |  |  |  |  |  |  |  |  |  |  |  |  |  |  |
| ... 15-29 | 1.00 |  |  |  |  | 1.00 |  |  |  |  | 0.00 |  |  |  |  |
| ... 30-49 | 1.60 | 1.41 | 1.81 | 0.000 | *** | 1.55 | 1.32 | 1.83 | 0.000 | *** | 0.40 | 0.07 | 0.73 | 0.017 | ** |
| ... 50+ | 1.56 | 1.34 | 1.81 | 0.000 | *** | 2.34 | 1.96 | 2.79 | 0.000 | *** | 0.29 | -0.06 | 0.64 | 0.104 |  |
| Marital status |  |  |  |  |  |  |  |  |  |  |  |  |  |  |  |
| ... married | 1.00 |  |  |  |  | 1.00 |  |  |  |  | 0.00 |  |  |  |  |
| ... other | 1.53 | 1.32 | 1.76 | 0.000 | *** | 1.41 | 1.18 | 1.68 | 0.000 | *** | -0.31 | -0.64 | 0.03 | 0.074 | * |
| Education |  |  |  |  |  |  |  |  |  |  |  |  |  |  |  |
| ... no education | 1.00 |  |  |  |  | 1.00 |  |  |  |  | 0.00 |  |  |  |  |
| ... up primary education (up to 7 years) | 1.18 | 1.04 | 1.35 | 0.013 | ** | 1.11 | 0.97 | 1.28 | 0.127 |  | 0.11 | -0.16 | 0.39 | 0.421 |  |
| ... secondary education and above (8 years and above) | 1.16 | 1.02 | 1.33 | 0.024 | ** | 0.91 | 0.79 | 1.05 | 0.208 |  | 0.09 | -0.20 | 0.38 | 0.552 |  |
| Employment |  |  |  |  |  |  |  |  |  |  |  |  |  |  |  |
| ... self-employed | 1.00 |  |  |  |  | 1.00 |  |  |  |  | 0.00 |  |  |  |  |
| ... salaried | 1.62 | 1.33 | 1.97 | 0.000 | *** | 1.15 | 0.94 | 1.40 | 0.166 |  | -0.07 | -0.47 | 0.33 | 0.725 |  |
| ... casual labour | 1.46 | 1.18 | 1.80 | 0.000 | *** | 1.17 | 0.94 | 1.46 | 0.157 |  | -0.03 | -0.46 | 0.40 | 0.891 |  |
| ... not working for employment | 1.04 | 0.90 | 1.19 | 0.594 |  | 1.08 | 0.93 | 1.26 | 0.323 |  | 0.32 | 0.03 | 0.62 | 0.032 | ** |
| Caste |  |  |  |  |  |  |  |  |  |  |  |  |  |  |  |
| ... SC and ST | 1.00 |  |  |  |  | 1.00 |  |  |  |  | 0.00 |  |  |  |  |
| ... OBC/ other/ no caste | 0.93 | 0.86 | 1.02 | 0.132 |  | 1.11 | 1.02 | 1.22 | 0.018 | ** | 0.09 | -0.09 | 0.26 | 0.336 |  |
| Religion |  |  |  |  |  |  |  |  |  |  |  |  |  |  |  |
| ... Hinduism | 1.00 |  |  |  |  | 1.00 |  |  |  |  | 0.00 |  |  |  |  |
| ... Others | 0.83 | 0.72 | 0.96 | 0.014 | ** | 1.06 | 0.90 | 1.24 | 0.466 |  | 0.07 | -0.25 | 0.40 | 0.663 |  |
| State |  |  |  |  |  |  |  |  |  |  |  |  |  |  |  |
| ... Bihar | 1.00 |  |  |  |  | 1.00 |  |  |  |  | 0.00 |  |  |  |  |
| ... Chhattisgarh | 0.48 | 0.42 | 0.54 | 0.000 | *** | 0.47 | 0.41 | 0.55 | 0.000 | *** | 0.87 | 0.59 | 1.16 | 0.000 | *** |
| ... Gujarat | 1.59 | 1.36 | 1.87 | 0.000 | *** | 0.40 | 0.34 | 0.46 | 0.000 | *** | -0.49 | -0.81 | -0.18 | 0.002 | *** |
| ... Meghalaya | 2.46 | 2.01 | 3.01 | 0.000 | *** | 0.09 | 0.07 | 0.12 | 0.000 | *** | 0.69 | 0.11 | 1.27 | 0.019 | ** |
| ... Tamil Nadu | 2.34 | 1.94 | 2.83 | 0.000 | *** | 0.43 | 0.37 | 0.51 | 0.000 | *** | 1.02 | 0.68 | 1.35 | 0.000 | *** |
| ... Kerala | 0.69 | 0.55 | 0.86 | 0.001 | *** | 0.39 | 0.30 | 0.51 | 0.000 | *** | 1.10 | 0.57 | 1.64 | 0.000 | *** |
| ... Uttar Pradesh | 1.42 | 1.26 | 1.61 | 0.000 | *** | 0.54 | 0.48 | 0.61 | 0.000 | *** | 0.40 | 0.17 | 0.64 | 0.001 | *** |
| Head of household's education |  |  |  |  |  |  |  |  |  |  |  |  |  |  |  |
| ... no education | 1.00 |  |  |  |  | 1.00 |  |  |  |  | 0.00 |  |  |  |  |
| ... up to primary education (up to 7 years) | 1.08 | 0.96 | 1.21 | 0.196 |  | 1.00 | 0.88 | 1.13 | 0.993 |  | 0.12 | -0.12 | 0.36 | 0.346 |  |
| ... secondary education and above (8 years and above) | 1.07 | 0.95 | 1.19 | 0.255 |  | 1.00 | 0.89 | 1.13 | 0.996 |  | -0.01 | -0.24 | 0.22 | 0.916 |  |
| Head of household's employment |  |  |  |  |  |  |  |  |  |  |  |  |  |  |  |
| ... self-employed | 1.00 |  |  |  |  | 1.00 |  |  |  |  | 0.00 |  |  |  |  |
| ... salaried | 0.85 | 0.73 | 0.99 | 0.031 | ** | 0.87 | 0.74 | 1.01 | 0.074 | * | 0.16 | -0.15 | 0.46 | 0.306 |  |
| ... casual labour | 0.78 | 0.69 | 0.88 | 0.000 | *** | 0.98 | 0.86 | 1.12 | 0.817 |  | 0.23 | -0.03 | 0.48 | 0.082 | * |
| ... not working for employment | 0.88 | 0.78 | 1.00 | 0.047 | ** | 0.96 | 0.84 | 1.09 | 0.526 |  | 0.36 | 0.11 | 0.61 | 0.005 | *** |
| Household wealth (quintiles) |  |  |  |  |  |  |  |  |  |  |  |  |  |  |  |
| ... Q1 (poorest) | 1.00 |  |  |  |  | 1.00 |  |  |  |  | 0.00 |  |  |  |  |
| ... Q2 | 1.23 | 1.09 | 1.39 | 0.001 | *** | 1.15 | 1.01 | 1.31 | 0.030 | ** | -0.01 | -0.26 | 0.24 | 0.931 |  |
| ... Q3 | 1.20 | 1.06 | 1.37 | 0.005 | *** | 1.18 | 1.03 | 1.35 | 0.019 | ** | -0.04 | -0.31 | 0.23 | 0.785 |  |
| ... Q4 | 1.36 | 1.19 | 1.56 | 0.000 | *** | 1.26 | 1.09 | 1.45 | 0.002 | *** | -0.04 | -0.33 | 0.25 | 0.775 |  |
| ... Q5 (least poor) | 1.33 | 1.15 | 1.54 | 0.000 | *** | 1.31 | 1.13 | 1.54 | 0.001 | *** | 0.02 | -0.30 | 0.33 | 0.910 |  |
| Household size | 0.96 | 0.95 | 0.98 | 0.000 | *** | 0.93 | 0.91 | 0.95 | 0.000 | *** | 0.04 | 0.00 | 0.08 | 0.077 | * |

Reference categories for the interaction verification and empowerment: no verification # no/low empowerment, no verification # medium empowerment, no verification # high empowerment, verification # no/low empowerment.
Abbreviations: AOR = Adjusted odds ratio, CI = Confidence interval, OOPE = out-of-pocket expenditure, SC + ST = scheduled castes and tribes, OBC = other backward classes.
\*Significant at 10%, \*\*Significant at 5%, \*\*\*Significant at 1%.

### Empowerment and outpatient service utilization

The bivariate results showed that highly empowered women were more likely to utilize outpatient services than other women (high empowerment: 15.4%, medium: 13.41%, no/ low: 11.71%, p < 0.001) (see Table 3, column 2). In addition, women who had undergone the verification process were more likely to use outpatient services than women with no verification (verification: 15.33%, no verification: 7.34%, p < 0.001).

However, in the multivariate logistic regressions (see Table 4, column 2), the association between empowerment and verification of outpatient service use was not found to be statistically significant. Looking at verification, women who had undergone the verification process had a statistically higher probability of utilizing outpatient services (AOR: 1.51, 95% CI: 1.27-1.80). The coefficient of the interaction term suggests that there was no difference in utilization by empowerment levels or by verification, meaning that empowerment did not moderate the association between verification in PM-JAY and outpatient service utilization.

### Empowerment and OOPE for outpatient services

The bivariate analysis in Table 3, column 3, showed significant differences in OOPE by women’s empowerment levels: women with no/low empowerment experienced higher OOPE (mean: 12,278 INR, SD: 24,805 INR) than women with medium empowerment (mean: 10,966 INR, SD: 24,274 INR). Women with high empowerment experienced the lowest OOPE (mean: 10,610 INR, SD: 20,660 INR).

These findings were also reflected in the GLM regression results of Table 4, column 3. We observed that women with medium empowerment experienced significantly lower OOPE for outpatient services than women with no/low empowerment (coef: -0.49, p = 0.020), whereas the result for women with high empowerment was not statistically significant. Similarly, women who had undergone the verification process incurred lower OOPE (coef = -0.37, p = 0.034). In contrast, the results of the interaction term indicate that women who were both medium empowered and verified in PM-JAY experienced higher OOPE (coef = 0.45, p = 0.058). Because our model included main effects and an interaction term, this finding reflects an additional effect of medium-empowered and verified women relative to the reference categories (no verification and no/low empowerment, no verification and medium empowerment, no verification and high empowerment, verification and no/low empowerment), rather than a direct comparison across groups.

## DISCUSSION

This study is, to our knowledge, the first to examine women’s empowerment and its association with verification in PM-JAY, outpatient service utilization, and resulting OOPE. Three main findings emerge: first, women’s empowerment was associated with higher odds of PM-JAY verification, but second, not with outpatient service utilization. Third, either medium levels of empowerment or PM-JAY verification were associated with lower OOPE for outpatient services, whereas women who were both verified in PM-JAY and medium empowered experienced higher OOPE. In the following, we discuss the findings for each outcome variable in the broader Indian and global context and outline their policy implications.

First, our findings show that women reporting high levels of empowerment were more likely to be verified than women with medium or no/ low empowerment. Although evidence specifically linking women’s empowerment to PFHI enrolment (and even less so, simplified verification or entitlement procedures) remains limited, these findings are consistent with broader Indian literature on gender and enrolment in PFHIs. For example, women’s enrolment into RSBY was shaped not only by eligibility criteria, but also by intra-households relations, with female-headed households showing higher enrolment rates in some cases[30–32]. Similar patterns have been observed internationally. A study across 29 sub-Saharan African countries found that women’s decision-making autonomy and agency were significantly associated with higher likelihood of health insurance enrolment and access to formal healthcare systems[12].

One possible explanation is that empowered women may have greater autonomy in seeking healthcare and navigating administrative steps required for insurance enrolment and verification. PM-JAY access requires interaction with digital systems, submission of documents, and engagement with healthcare institutions, all of which may disadvantage women with limited mobility, lower literacy, or restricted decision-making power[25,47]. Existing evidence from India suggests that awareness and understanding of PM-JAY remain unevenly distributed across populations, with women exhibiting lower levels of scheme awareness and engagement[28,29]. Inequalities may therefore emerge even before healthcare services are utilized, at the verification stage itself. Since verification functions as a gateway to PM-JAY benefits, disparities at this stage may contribute to unequal access to covered services despite formal eligibility. Simplified verification procedures, including a single verification process for all public benefits schemes using existing channels like the national digital identifier AADHAAR, community-based awareness initiatives targeted at women, and assistance with documentation, may help improve equitable access to PM-JAY. In these processes, Ayushman Mitras, Accredited Social Health Activists (ASHA workers), and hospital-based frontline staff likely play an important role, as shown in prior studies[48–50]. Evidence from other countries similarly suggests that outreach tailored to women’s needs and practical enrolment support can improve equitable insurance participation[51–53].

Second, although the bivariate analysis showed that women with either high empowerment levels or PM-JAY verification were more likely to use outpatient services, only verification remained significant in the multivariate model. Additionally, we did not observe that women who were both empowered and verified under PM-JAY used outpatient services more than women with no/low empowerment and no verification (in comparison to women with no verification and no/low empowerment, no verification and medium empowerment, no verification and high empowerment, verification and no/low empowerment). This suggests that outpatient service utilization may be driven more by more egalitarian integration into formal healthcare systems through PM-JAY verification alone, rather than through differential effects of verification across women with different levels of empowerment. Again, this may reflect the fact that verification of women into PM-JAY is reflective of their broader engagement with healthcare providers and administrative systems, which could facilitate subsequent healthcare-seeking behavior. This is consistent with evidence from the international literature showing that administrative contact and navigation support on the one hand, and empowerment on the other hand, can improve service use[5,13,14,48,54].

The finding that PM-JAY verification was associated with higher outpatient service utilization despite outpatient care not being formally covered under the scheme is noteworthy. This is indicative of possible income and substitution effects due to health insurance coverage[8]. In PFHI schemes where both inpatient and outpatient services are covered with varying levels of cost-sharing, substitution and complementary effects may be observed: e.g., in China, recent evidence shows that patients in areas with higher cost-sharing for outpatient services used more inpatient services[55]. These effects need to be substantiated by further rigorous, causal research. These findings may also reflect indirect effects of insurance participation, including greater familiarity with healthcare institutions, increased trust in formal healthcare services, or improved awareness of available care options. Evidence from India suggests that PFHI can generate spillover effects beyond the services directly covered by the scheme. For example, RSBY was associated with increased use of outpatient services not covered by the insurance, and for childhood immunizations[56,57]. A systematic review of spillover effects of health insurance in low- and middle income countries (LMIC) found that these effects are most consistently observed for outcomes related to reduced disease transmission, particularly vaccination and mass drug administration[58]. For PM-JAY, further research is needed to better understand the mechanisms underlying this association.

Third, among women who used outpatient care and incurred positive spending, those with either medium levels of empowerment or PM-JAY verification were associated with lower OOPE than women with no/low empowerment and no PM-JAY verification. One possible explanation is that empowerment may provide women with sufficient autonomy and healthcare awareness to seek timely care and use lower-cost services, while PM-JAY verification may reflect greater familiarity with healthcare institutions and available support systems rather than direct insurance-related financial protection. Because outpatient services are not covered under PM-JAY, lower OOPE among verified women likely reflects improved awareness, for instance through engagement with frontline workers, ASHAs and Ayushman Mitras, as well as better navigation of the healthcare system.

Interestingly, OOPE was higher among women with both PM-JAY verification and medium levels of empowerment, indicating that the association between verification and OOPE differed for women with medium empowerment relative to women with no/low empowerment and no PM-JAY verification. This finding may indicate that women who are moderately empowered and successfully integrated into PM-JAY are more likely to actively seek healthcare services, including diagnostics, medicines, and follow-up services that remain outside PM-JAY coverage, or that they seek services that are qualitatively different from women in other empowerment and verification strata, e.g., from more expensive private providers or of higher quality. In this context, increased healthcare utilization may translate into higher short-term expenditure despite better healthcare access. Similar patterns have been reported in India and other settings in the Global South, where improved healthcare access and insurance enrolment were associated with increased utilization of uncovered or partially covered services, particularly outpatient care and medicines[59–61]. These findings may therefore reflect previously unmet healthcare needs being addressed among women who are sufficiently empowered to seek care and sufficiently connected to the healthcare system through PM-JAY verification.

From a policy perspective, our findings highlight important limitations in the current scope of financial protection under PM-JAY and in broader health system reforms in India. Although PM-JAY has substantially expanded access to inpatient services, outpatient care and medicines continue to account for a large share of OOPE among Indian households[62].

Given that this study specifically examined the association between women’s empowerment, PM-JAY verification, outpatient service utilization, and OOPE, the findings suggest that equitable healthcare access depends not only on insurance coverage itself, but also on women’s ability to engage with healthcare systems and navigate care pathways. Recent studies from India and elsewhere emphasize the need to strengthen primary healthcare, outpatient services, and medicine coverage alongside insurance-based reforms to achieve meaningful financial protection[63–65]. Similar conclusions have been reported in other LMIC settings, where health insurance schemes alone were insufficient to eliminate healthcare-related financial hardship without parallel investments in accessible primary care and gender-sensitive outreach systems[66–68]. Future reforms under PM-JAY may therefore benefit from stronger integration with primary healthcare services, expanded outpatient coverage, and continued investment in community-based awareness and support mechanisms targeting socially disadvantaged women.

### Methodological considerations

This study has several additional limitations that should be considered when interpreting the findings. First, the analysis provides observational evidence on the associations between women’s empowerment, PM-JAY verification, outpatient service utilization, and OOPE, but the study design does not permit causal inference. Given the rapid nationwide implementation of PM-JAY, it was not possible to establish a suitable comparison group to assess causal effects. Second, although the survey included data from six socio-economically and geographically diverse Indian states, the findings cannot be generalized to settings that are different from the study states. Third, the sampling frame relied on PM-JAY eligibility lists derived largely from the SECC 2011 database. As many household records contained outdated or incomplete contact information, locating eligible households was challenging, and data collection had to be restricted primarily to rural and semi-urban areas. Consequently, households from larger urban settings were not included in the analysis, limiting the generalizability of the findings to urban populations. However, this limitation may be less pronounced given that a substantial proportion of PM-JAY eligible households reside in rural areas. Fourth, the study relied on self-reported data regarding healthcare utilization, PM-JAY verification, and expenditure, which may be subject to recall bias and reporting inaccuracies.

## CONCLUSION

Our study suggests that women’s empowerment plays an important role in their ability to navigate insurance systems, access care, and benefit from financial protection. At the same time, the findings show that empowerment alone is not sufficient: equitable access depends on gender-sensitive, responsive health systems that reduce administrative barriers, strengthen frontline support, and better connect insurance with primary and outpatient care. For PM-JAY and similar PFHI schemes, achieving UHC will therefore require more than expanding eligibility; it will require ensuring that women can meaningfully engage with the system and translate entitlement into timely, affordable care.

## Data Availability

All data produced in the present study are available upon reasonable request to the principal investigator Manuela De Allegri.

## Ethics approval and consent to participate

### Patient consent for publication

Not applicable.

### Ethics approval

We obtained ethical approval from the Ethics Commission of the Medical Faculty via approval number 2019-05-09 ZB 25519, Heidelberg University, Germany, and from Sigma Institutional Review Board, New Delhi, India, via approval number 10057/IRB/19–20. All respondents provided written (or, when illiterate, with thumbprint) informed consent to participate in the study. The research study adhered to the Declaration of Helsinki.

## Data sharing statement

Proposals for data access should be directed to the Principal Investigator (MDA); data requestors will need to sign a data access agreement. The proposal for the use of data will be reviewed by a committee identified by MDA and the study funder.

## Author contributions

SS, SZ, and MDA defined the research question and the corresponding study design. SS and SZ conducted the analyses, interpreted the data, and wrote the first draft of the manuscript. All authors contributed to finalizing the draft and have approved the paper.

## Acknowledgments

The authors would like to thank the enumerators and respondents for their cooperation and contribution to the study.

