## Supplementary File 1 for "Women’s empowerment and publicly-funded health insurance in India: associations with PM-JAY verification, outpatient service use, and out-of-pocket spending across six states"

**Supplementary File 1: Construction of Women's Empowerment Index**

**Exploratory Factor Analysis, using 18 Items:**

Decision making: women_spend_dec women_healt_dec women_ownhealt_dec women_seekhealt_dec

Permission to go to places: women_places_mrkt women_places_hosp women_places_friend women_places_out

Perceived problems: women_treat_permis women_treat_money women_treat_dist women_treat_transp women_treat_comp women_treat_nofem

Financial empowerment: women_money women_saving women_selfhelp women_micro

All items coded 0 (no or low empowerment) or 1 (high empowerment)

**Summary Statistics**

| Variable | Mean | SD | Skewness | Kurtosis |
|---|---|---|---|---|
| women_spen~c | .5104326 | .4999085 | -.0417395 | 1.001742 |
| women_heal~c | .5203784 | .4996019 | -.0815812 | 1.006655 |
| women_ownh~c | .5498679 | .4975243 | -.200471 | 1.040189 |
| women_seek~c | .5560579 | .4968649 | -.2256542 | 1.05092 |
| women_pla~kt | .6224092 | .4848013 | -.5050048 | 1.25503 |
| women_plac~p | .5110586 | .4998951 | -.0442451 | 1.001958 |
| women_plac~d | .6132285 | .4870275 | -.4649942 | 1.21622 |
| women_pla~ut | .5201002 | .4996132 | -.0804657 | 1.006475 |
| women_trea~s | .5327584 | .4989431 | -.1313157 | 1.017244 |
| women_trea~y | .200306 | .4002433 | 1.497611 | 3.242839 |
| women_trea~t | .2092085 | .4067577 | 1.429852 | 3.044476 |
| women_tre~sp | .2414105 | .4279535 | 1.208536 | 2.460558 |
| women_tre~mp | .364237 | .4812323 | .5642501 | 1.318378 |
| women_trea~m | .3653498 | .4815449 | .5592619 | 1.312774 |
| women_money | .6184448 | .4857852 | -.4876599 | 1.237812 |
| women_saving | .7412714 | .4379514 | -1.101856 | 2.214087 |
| women_self~p | .2192934 | .4137822 | 1.356832 | 2.840992 |
| women_micro | .1011963 | .3015989 | 2.644688 | 7.994377 |

**Correlation matrix**

| | wo~d_dec | women_h~ | w~wnhe~c | w~ekhe~c | women~kt | wome~osp | women_~d | women~ut | women_~s |
|---|---|---|---|---|---|---|---|---|---|
| women_spen~c | 1.0000 | | | | | | | | |
| women_heal~c | 0.7867 | 1.0000 | | | | | | | |
| women_ownh~c | 0.6719 | 0.7703 | 1.0000 | | | | | | |
| women_seek~c | 0.6029 | 0.7102 | 0.8165 | 1.0000 | | | | | |
| women_pla~kt | 0.2159 | 0.2269 | 0.2507 | 0.2188 | 1.0000 | | | | |
| women_plac~p | 0.1835 | 0.2103 | 0.2144 | 0.2101 | 0.6407 | 1.0000 | | | |
| women_plac~d | 0.1718 | 0.1855 | 0.2081 | 0.2087 | 0.5963 | 0.6102 | 1.0000 | | |
| women_pla~ut | 0.1980 | 0.2028 | 0.1850 | 0.1840 | 0.5693 | 0.6323 | 0.6341 | 1.0000 | |
| women_trea~s | 0.0700 | 0.1169 | 0.1286 | 0.1390 | 0.0959 | 0.0681 | 0.1161 | 0.0388 | 1.0000 |
| women_trea~y | 0.0073 | 0.0349 | 0.0749 | 0.0915 | 0.1066 | 0.0658 | 0.1402 | 0.0623 | 0.3656 |
| women_trea~t | 0.0320 | 0.0475 | 0.0739 | 0.0703 | 0.0744 | 0.0663 | 0.1132 | 0.0361 | 0.3371 |
| women_tre~sp | -0.0025 | 0.0295 | 0.0563 | 0.0569 | 0.0555 | 0.0358 | 0.1140 | 0.0302 | 0.3198 |
| women_tre~mp | 0.0188 | 0.0595 | 0.0861 | 0.0855 | 0.0678 | 0.0444 | 0.1260 | 0.0209 | 0.4420 |
| women_trea~m | 0.0539 | 0.0822 | 0.1189 | 0.1172 | 0.0755 | 0.0819 | 0.1156 | 0.0688 | 0.3742 |
| women_money | 0.1988 | 0.2232 | 0.2321 | 0.2122 | 0.2479 | 0.2015 | 0.1773 | 0.2205 | 0.0935 |
| women_saving | 0.1286 | 0.1373 | 0.1266 | 0.0890 | 0.1868 | 0.1687 | 0.1709 | 0.1989 | 0.0413 |
| women_self~p | -0.0412 | -0.0278 | -0.0225 | -0.0248 | 0.0824 | 0.0641 | 0.0433 | 0.0606 | -0.0110 |
| women_micro | -0.0095 | -0.0015 | 0.0143 | 0.0074 | 0.1029 | 0.1021 | 0.0856 | 0.1183 | -0.0750 |

| | ~t_money | women~st | wome~nsp | women~mp | women_~m | ~n_money | women_~g | women~lp | women_~o |
|---|---|---|---|---|---|---|---|---|---|
| women_trea~y | 1.0000 | | | | | | | | |
| women_trea~t | 0.5697 | 1.0000 | | | | | | | |
| women_tre~sp | 0.5262 | 0.6696 | 1.0000 | | | | | | |
| women_tre~mp | 0.4734 | 0.5282 | 0.5400 | 1.0000 | | | | | |
| women_trea~m | 0.3785 | 0.4425 | 0.4040 | 0.4561 | 1.0000 | | | | |
| women_money | 0.0658 | 0.0911 | 0.0707 | 0.0974 | 0.0715 | 1.0000 | | | |
| women_saving | 0.0247 | 0.0466 | 0.0549 | 0.0927 | 0.0733 | 0.3906 | 1.0000 | | |
| women_self~p | -0.0422 | -0.0197 | -0.0083 | -0.0239 | -0.0112 | -0.0104 | 0.0698 | 1.0000 | |
| women_micro | -0.0654 | -0.0303 | -0.0260 | -0.0230 | -0.0036 | 0.0632 | 0.1034 | 0.4425 | 1.0000 |

**FA to determine number of factors**

| | |
|---|---|
| Factor analysis/correlation | Number of obs = 14,378 |
| Method: principal-component factors | Retained factors = 5 |
| Rotation: (unrotated) | Number of params = 80 |

| Factor | Eigenvalue | Difference | Proportion | Cumulative |
|---|---|---|---|---|
| Factor1 | 4.33807 | 1.27718 | 0.2410 | 0.2410 |
| Factor2 | 3.06089 | 0.81055 | 0.1700 | 0.4111 |
| Factor3 | 2.25034 | 0.85407 | 0.1250 | 0.5361 |
| Factor4 | 1.39627 | 0.22761 | 0.0776 | 0.6136 |
| Factor5 | 1.16866 | 0.37752 | 0.0649 | 0.6786 |
| Factor6 | 0.79113 | 0.14562 | 0.0440 | 0.7225 |
| Factor7 | 0.64552 | 0.04034 | 0.0359 | 0.7584 |
| Factor8 | 0.60518 | 0.06530 | 0.0336 | 0.7920 |
| Factor9 | 0.53988 | 0.03507 | 0.0300 | 0.8220 |
| Factor10 | 0.50481 | 0.04108 | 0.0280 | 0.8500 |
| Factor11 | 0.46372 | 0.02845 | 0.0258 | 0.8758 |
| Factor12 | 0.43527 | 0.00516 | 0.0242 | 0.9000 |
| Factor13 | 0.43011 | 0.06119 | 0.0239 | 0.9239 |
| Factor14 | 0.36892 | 0.03875 | 0.0205 | 0.9444 |
| Factor15 | 0.33017 | 0.01969 | 0.0183 | 0.9627 |
| Factor16 | 0.31048 | 0.11910 | 0.0172 | 0.9800 |
| Factor17 | 0.19137 | 0.02215 | 0.0106 | 0.9906 |
| Factor18 | 0.16922 | . | 0.0094 | 1.0000 |

LR test: independent vs. saturated: chi2(153) = 1.1e+05 Prob>chi2 = 0.0000

Factor loadings (pattern matrix) and unique variances

| Variable | Factor1 | Factor2 | Factor3 | Factor4 | Factor5 | Uniqueness |
|---|---|---|---|---|---|---|
| women_spen~c | 0.6137 | -0.3699 | -0.4649 | 0.0309 | -0.0465 | 0.2672 |
| women_heal~c | 0.6772 | -0.3571 | -0.4977 | 0.0555 | -0.0591 | 0.1596 |
| women_ownh~c | 0.6987 | -0.3180 | -0.4878 | 0.0691 | -0.0818 | 0.1612 |
| women_seek~c | 0.6705 | -0.2925 | -0.4723 | 0.0527 | -0.1129 | 0.2263 |
| women_pla~kt | 0.6029 | -0.2203 | 0.4998 | -0.1423 | -0.0771 | 0.3120 |
| women_plac~p | 0.5807 | -0.2387 | 0.5414 | -0.1843 | -0.1227 | 0.2637 |
| women_plac~d | 0.5998 | -0.1403 | 0.5297 | -0.2015 | -0.1347 | 0.2812 |
| women_pla~ut | 0.5590 | -0.2550 | 0.5497 | -0.1668 | -0.0701 | 0.2876 |
| women_trea~s | 0.3785 | 0.4770 | -0.0826 | -0.0140 | -0.0217 | 0.6218 |
| women_trea~y | 0.3890 | 0.6460 | -0.0026 | -0.0432 | -0.0684 | 0.4248 |
| women_trea~t | 0.4067 | 0.7062 | -0.0211 | 0.0434 | -0.0323 | 0.3325 |
| women_tre~sp | 0.3753 | 0.7033 | -0.0119 | 0.0593 | -0.0268 | 0.3602 |
| women_tre~mp | 0.4009 | 0.6676 | -0.0365 | 0.0619 | 0.0318 | 0.3874 |
| women_trea~m | 0.3915 | 0.5439 | -0.0371 | 0.0644 | -0.0328 | 0.5443 |
| women_money | 0.4170 | -0.1007 | 0.0789 | 0.1283 | 0.6880 | 0.3199 |
| women_saving | 0.3135 | -0.0817 | 0.1763 | 0.2434 | 0.7249 | 0.2792 |
| women_self~p | 0.0217 | -0.0744 | 0.2635 | 0.7668 | -0.2529 | 0.2725 |
| women_micro | 0.0686 | -0.1332 | 0.2878 | 0.7635 | -0.1663 | 0.2841 |

Scree plot:

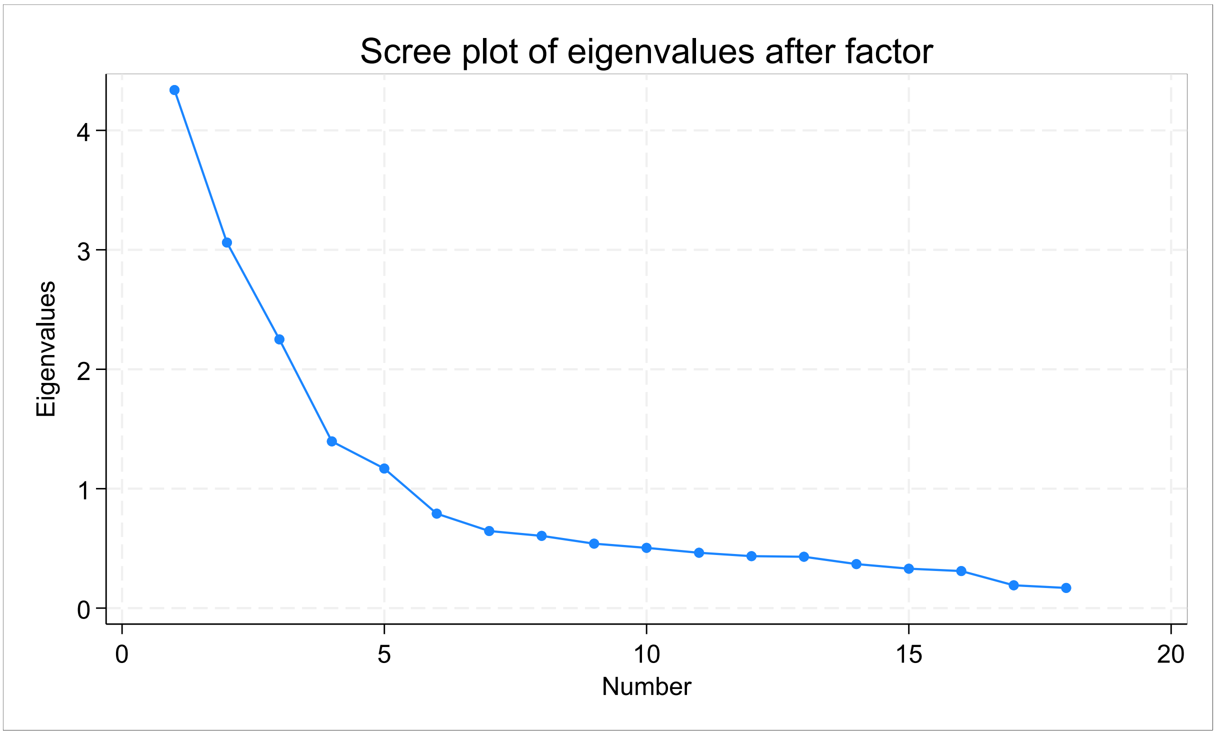

**FA analysis using maximum-likelihood and factors based on scree plot**

4 factors (unrotated solution):

| | | | |
|---|---|---|---|
| Factor analysis/correlation | Number of obs | = | 14,378 |
| Method: maximum likelihood | Retained factors | = | 4 |
| Rotation: (unrotated) | Number of params | = | 66 |
| | Schwarz's BIC | = | 7992.43 |
| Log likelihood = -3680.289 | (Akaike's) AIC | = | 7492.58 |

| Factor | Eigenvalue | Difference | Proportion | Cumulative |
|---|---|---|---|---|
| Factor1 | 3.73812 | 1.12300 | 0.3983 | 0.3983 |
| Factor2 | 2.61512 | 0.52123 | 0.2786 | 0.6769 |
| Factor3 | 2.09389 | 1.15489 | 0.2231 | 0.9000 |
| Factor4 | 0.93900 | . | 0.1000 | 1.0000 |

LR test: independent vs. saturated: chi2(153) = 1.1e+05 Prob>chi2 = 0.0000

LR test: 4 factors vs. saturated: chi2(87) = 7355.71 Prob>chi2 = 0.0000

Factor loadings (pattern matrix) and unique variances

| Variable | Factor1 | Factor2 | Factor3 | Factor4 | Uniqueness |
|---|---|---|---|---|---|

| | | | | | |
|---|---|---|---|---|---|
| women_spen~c | 0.7478 | -0.2296 | -0.1097 | -0.0082 | 0.3760 |
| women_heal~c | 0.8356 | -0.2281 | -0.1473 | 0.0131 | 0.2280 |
| women_ownh~c | 0.8631 | -0.1957 | -0.1694 | 0.0411 | 0.1863 |
| women_seek~c | 0.8120 | -0.1725 | -0.1619 | 0.0308 | 0.2837 |
| women_pla~kt | 0.4552 | 0.2962 | 0.5311 | -0.1032 | 0.4123 |
| women_plac~p | 0.4385 | 0.3030 | 0.5900 | -0.1303 | 0.3508 |
| women_plac~d | 0.4291 | 0.3726 | 0.5226 | -0.1318 | 0.3866 |
| women_pla~ut | 0.4146 | 0.2827 | 0.5942 | -0.1134 | 0.3823 |
| women_trea~s | 0.1998 | 0.3954 | -0.2379 | -0.0224 | 0.7467 |
| women_trea~y | 0.1626 | 0.6032 | -0.3013 | -0.0043 | 0.5189 |
| women_trea~t | 0.1702 | 0.6868 | -0.3733 | 0.0662 | 0.3556 |
| women_tre~sp | 0.1433 | 0.6694 | -0.3666 | 0.0749 | 0.3913 |
| women_tre~mp | 0.1689 | 0.5938 | -0.3321 | 0.0604 | 0.5050 |
| women_trea~m | 0.1908 | 0.4775 | -0.2492 | 0.0565 | 0.6703 |
| women_money | 0.3116 | 0.1006 | 0.0967 | 0.0240 | 0.8828 |
| women_saving | 0.2026 | 0.1057 | 0.1372 | 0.0732 | 0.9236 |
| women_self~p | -0.0025 | 0.0350 | 0.1953 | 0.4853 | 0.7251 |
| women_micro | 0.0457 | 0.0302 | 0.2984 | 0.7871 | 0.2885 |

Oblique (promax) rotation (as factors are expected to be correlated)

| | | | | |
|---|---|---|---|---|
| Factor analysis/correlation | | Number of obs | = | 14,378 |
| Method: maximum likelihood | | Retained factors | = | 4 |
| Rotation: oblique promax (Kaiser | off) | Number of params | = | 66 |
| | | Schwarz's BIC | = | 7992.43 |
| Log likelihood = - 3680.289 | | (Akaike's) AIC | = | 7492.58 |

| | | | | | | |
|---|---|---|---|---|---|---|
| Factor | Variance | Proportion | Rotated | factors | are | correlated |
| Factor1 | 3.32440 | 0.3542 | | | | |
| Factor2 | 2.99081 | 0.3186 | | | | |
| Factor3 | 2.92009 | 0.3111 | | | | |
| Factor4 | 1.10438 | 0.1177 | | | | |

LR test: independent vs. saturated: chi2(153) = 1.1e+05 Prob>chi2 = 0.0000

LR test: 4 factors vs. saturated: chi2(87) = 7355.71 Prob>chi2 = 0.0000

Rotated factor loadings (pattern matrix) and unique variances

| | | | | | |
|---|---|---|---|---|---|
| Variable | Factor1 | Factor2 | Factor3 | Factor4 | Uniqueness |
| women_spen~c | 0.7946 | 0.0045 | -0.0672 | -0.0236 | 0.3760 |
| women_heal~c | 0.8870 | -0.0137 | -0.0340 | -0.0080 | 0.2280 |
| women_ownh~c | 0.9077 | -0.0235 | 0.0127 | 0.0171 | 0.1863 |
| women_seek~c | 0.8484 | -0.0162 | 0.0218 | 0.0079 | 0.2837 |
| women_pla~kt | 0.0269 | 0.7576 | -0.0060 | 0.0030 | 0.4123 |
| women_plac~p | -0.0176 | 0.8191 | -0.0377 | -0.0136 | 0.3508 |
| women_plac~d | -0.0323 | 0.7870 | 0.0561 | -0.0263 | 0.3866 |
| women_pla~ut | -0.0280 | 0.8012 | -0.0566 | 0.0035 | 0.3823 |
| women_trea~s | 0.0754 | 0.0181 | 0.4774 | -0.0567 | 0.7467 |
| women_trea~y | -0.0278 | 0.0333 | 0.6872 | -0.0470 | 0.5189 |
| women_trea~t | -0.0233 | -0.0212 | 0.8092 | 0.0124 | 0.3556 |
| women_tre~sp | -0.0397 | -0.0331 | 0.7898 | 0.0218 | 0.3913 |
| women_tre~mp | 0.0026 | -0.0222 | 0.7072 | 0.0125 | 0.5050 |
| women_trea~m | 0.0434 | 0.0093 | 0.5658 | 0.0218 | 0.6703 |
| women_money | 0.1850 | 0.2008 | 0.0675 | 0.0471 | 0.8828 |
| women_saving | 0.0792 | 0.1907 | 0.0483 | 0.1028 | 0.9236 |
| women_self~p | -0.0332 | -0.0052 | 0.0081 | 0.5247 | 0.7251 |
| women_micro | 0.0088 | -0.0229 | 0.0050 | 0.8477 | 0.2885 |

Factor rotation matrix

| | | | | |
|---|---|---|---|---|
| | Factor1 | Factor2 | Factor3 | Factor4 |
| Factor1 | 0.9609 | 0.5686 | 0.2520 | 0.0579 |
| Factor2 | -0.1693 | 0.7020 | -0.4536 | 0.3754 |
| Factor3 | -0.2178 | 0.4052 | 0.8524 | 0.0438 |
| Factor4 | 0.0234 | -0.1402 | 0.0646 | 0.9240 |

**Confirmatory Factor Analysis with four factor solution**

SEM Model

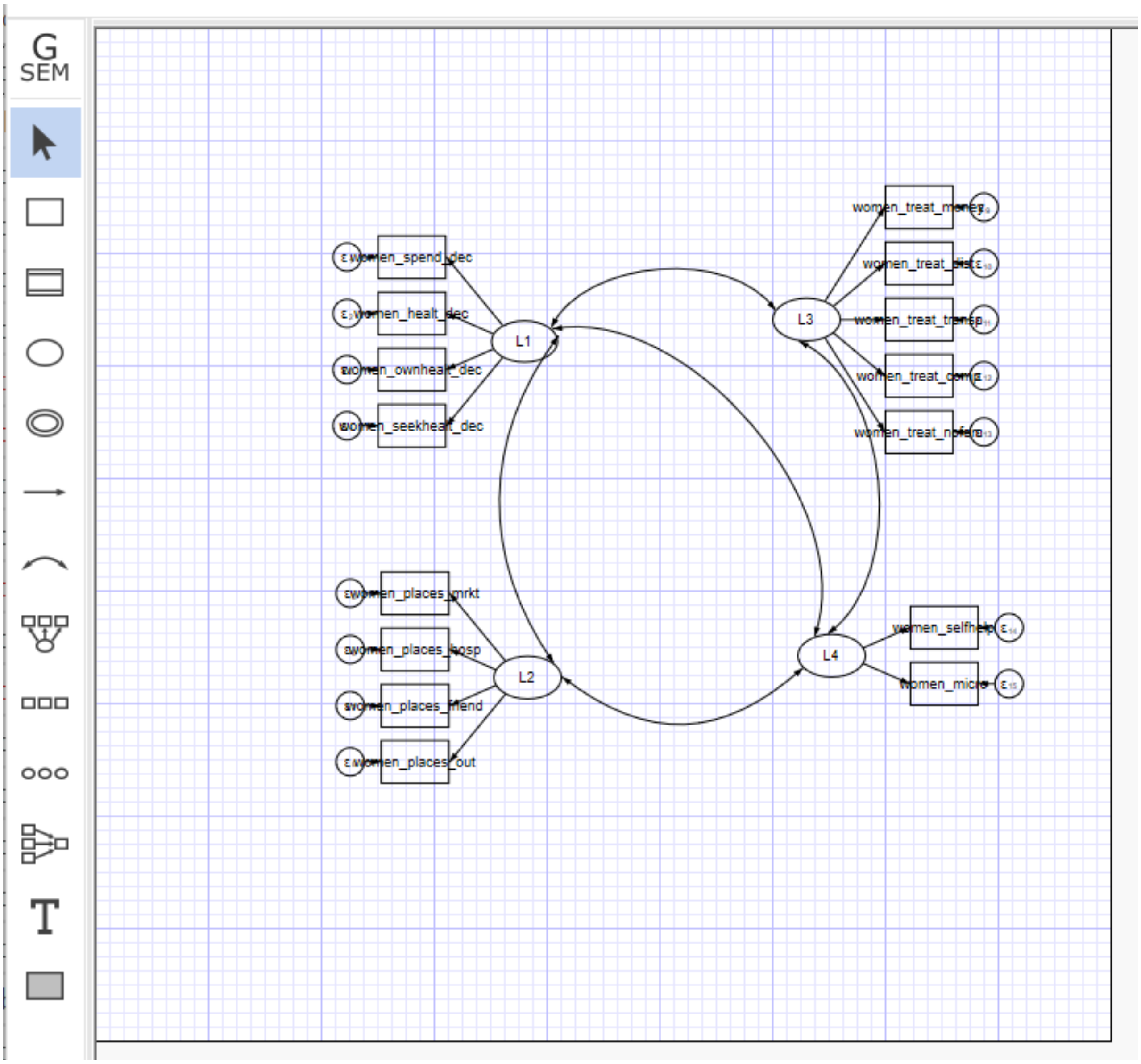

Endogenous variables

Measurement: women_spend_dec women_healt_dec women_ownhealt_dec women_seekhealt_dec women_places_mrkt women_places_hosp women_places_friend women_places_out women_treat_money

women_treat_dist women_treat_transp women_treat_comp women_treat_nofem women_selfhelp women_micro

Exogenous variables

Latent: L1 L2 L3 L4

Fitting target model:

Iteration 0:  Log likelihood = -88050.803
Iteration 1:  Log likelihood = -87584.494
Iteration 2:  Log likelihood = -87469.292  (not concave)
Iteration 3:  Log likelihood = -87467.612
Iteration 4:  Log likelihood =     -87467
Iteration 5:  Log likelihood = -87466.994
Iteration 6:  Log likelihood = -87466.994

Structural equation model                Number of obs = 14,378

Estimation method: ml

Log likelihood = -87466.994

( 1)  [women_spend_dec]L1 = 1
( 2)  [women_places_mrkt]L2 = 1
( 3)  [women_treat_money]L3 = 1
( 4)  [women_selfhelp]L4 = 1

| | | OIM | | | | |
|---|---|---|---|---|---|---|
| Standardized | Coefficient | std. err. | z | P>z | [95% conf. interval] | |
| Measurement | | | | | | |
| women_spend_dec | | | | | | |
| L1 | .7855776 | .0039751 | 197.62 | 0.000 | .7777865 | .7933686 |
| _cons | 1.021088 | .0102863 | 99.27 | 0.000 | 1.000927 | 1.041248 |
| women_healt_dec | | | | | | |
| L1 | .8764339 | .0028892 | 303.35 | 0.000 | .8707712 | .8820966 |
| _cons | 1.041622 | .0103577 | 100.57 | 0.000 | 1.021322 | 1.061923 |
| women_ownhealt_dec | | | | | | |
| L1 | .9026379 | .0025189 | 358.35 | 0.000 | .897701 | .9075748 |
| _cons | 1.105247 | .0105845 | 104.42 | 0.000 | 1.084501 | 1.125992 |
| women_seekhealt_dec | | | | | | |
| L1 | .847634 | .0030542 | 277.53 | 0.000 | .8416479 | .8536201 |
| _cons | 1.119172 | .0106352 | 105.23 | 0.000 | 1.098327 | 1.140017 |
| women_places_mrkt | | | | | | |
| L2 | .7672746 | .0043243 | 177.43 | 0.000 | .7587991 | .7757501 |
| _cons | 1.283889 | .0112638 | 113.98 | 0.000 | 1.261812 | 1.305965 |
| women_places_hosp | | | | | | |
| L2 | .8078285 | .0039102 | 206.59 | 0.000 | .8001646 | .8154924 |
| _cons | 1.022367 | .0102907 | 99.35 | 0.000 | 1.002198 | 1.042537 |

| |
|---|
| women_places_friend |
| L2 .7806083 .0042008 185.82 0.000 .7723749 .7888417 |
| _cons 1.259169 .0111663 112.76 0.000 1.237283 1.281054 |
| women_places_out |
| L2 .7788105 .0042021 185.34 0.000 .7705746 .7870465 |
| _cons 1.041042 .0103556 100.53 0.000 1.020745 1.061339 |
| women_treat_money |
| L3 .6846814 .005287 129.50 0.000 .674319 .6950437 |
| _cons .5004781 .0088465 56.57 0.000 .4831392 .517817 |
| women_treat_dist |
| L3 .8216167 .0039222 209.48 0.000 .8139293 .8293041 |
| _cons .5143499 .0088742 57.96 0.000 .4969569 .531743 |
| women_treat_transp |
| L3 .7910922 .004179 189.30 0.000 .7829015 .7992829 |
| _cons .5641241 .0089787 62.83 0.000 .5465261 .5817221 |
| women_treat_comp |
| L3 .6802188 .0053839 126.34 0.000 .6696665 .6907712 |
| _cons .7569103 .0094591 80.02 0.000 .7383709 .7754498 |
| women_treat_nofem |
| L3 .554263 .0065655 84.42 0.000 .5413949 .5671311 |
| _cons .75873 .0094641 80.17 0.000 .7401806 .7772794 |
| women_selfhelp |
| L4 .5494605 .024674 22.27 0.000 .5011004 .5978207 |
| _cons .5299914 .0089061 59.51 0.000 .5125357 .547447 |
| women_micro |
| L4 .8053318 .035265 22.84 0.000 .7362135 .87445 |
| _cons .3355443 .0085712 39.15 0.000 .318745 .3523436 |
| var(e.women_spend_dec) .3828679 .0062455 .3708206 .3953067 |
| var(e.women_healt_dec) .2318636 .0050644 .2221471 .2420051 |
| var(e.women_ownhealt_dec) .1852448 .0045473 .1765433 .1943752 |
| var(e.women_seekhealt_dec) .2815167 .0051777 .2715494 .2918498 |
| var(e.women_places_mrkt) .4112897 .0066359 .3984871 .4245036 |
| var(e.women_places_hosp) .3474131 .0063176 .3352489 .3600187 |
| var(e.women_places_friend) .3906507 .0065583 .3780058 .4037186 |
| var(e.women_places_out) .3934541 .0065453 .3808325 .4064941 |
| var(e.women_treat_money) .5312114 .0072398 .5172095 .5455925 |
| var(e.women_treat_dist) .324946 .0064451 .3125562 .337827 |
| var(e.women_treat_transp) .3741731 .006612 .3614357 .3873594 |
| var(e.women_treat_comp) .5373023 .0073245 .5231366 .5518516 |

| | | | | | | |
|---|---|---|---|---|---|---|
| var(e.women_treat_nofem) | .6927925 | .007278 | | | .6786737 | .707205 |
| var(e.women_selfhelp) | .6980931 | .0271148 | | | .6469216 | .7533123 |
| var(e.women_micro) | .3514408 | .0568001 | | | .2560238 | .4824185 |
| var(L1) | 1 | . | . | . | | |
| var(L2) | 1 | . | . | . | | |
| var(L3) | 1 | . | . | . | | |
| var(L4) | 1 | . | . | . | | |
| cov(L1,L2) | .3040347 | .0085772 | 35.45 | 0.000 | .2872238 | .3208457 |
| cov(L1,L3) | .0924978 | .0093318 | 9.91 | 0.000 | .0742079 | .1107877 |
| cov(L1,L4) | -.005204 | .0108331 | -0.48 | 0.631 | -.0264366 | .0160285 |
| cov(L2,L3) | .1232136 | .0095872 | 12.85 | 0.000 | .1044229 | .1420042 |
| cov(L2,L4) | .1584201 | .0111413 | 14.22 | 0.000 | .1365836 | .1802566 |
| cov(L3,L4) | -.0513538 | .0108718 | -4.72 | 0.000 | -.0726622 | -.0300454 |

LR test of model vs. saturated: chi2(84) = 4979.09 Prob > chi2 = 0.0000

**Final Score items:** alpha women_spend_dec women_healt_dec women_ownhealt_dec women_seekhealt_dec women_places_mrkt women_places_hosp women_places_friend women_places_out women_treat_money women_treat_dist women_treat_transp women_treat_comp women_treat_nofem women_selfhelp women_micro

Test scale = mean(unstandardized items)

Average interitem covariance: .0422044

Number of items in the scale: 15

Scale reliability coefficient: 0.7872
